# Comparison of Rapid Antigen Tests’ Performance between Delta (B.1.61.7; AY.X) and Omicron (B.1.1.529; BA1) Variants of SARS-CoV-2: Secondary Analysis from a Serial Home Self-Testing Study

**DOI:** 10.1101/2022.02.27.22271090

**Authors:** Apurv Soni, Carly Herbert, Andreas Filippaios, John Broach, Andres Colubri, Nisha Fahey, Kelsey Woods, Janvi Nanavati, Colton Wright, Taylor Orwig, Karen Gilliam, Vik Kheterpal, Thejas Suvarna, Chris Nowak, Summer Schrader, Honghuang Lin, Laurel O’Connor, Caitlin Pretz, Didem Ayturk, Elizabeth Orvek, Julie Flahive, Peter Lazar, Qiming Shi, Chad Achenbach, Robert Murphy, Matthew Robinson, Laura Gibson, Pamela Stamegna, Nathaniel Hafer, Katherine Luzuriaga, Bruce Barton, William Heetderks, Yukari C. Manabe, David McManus, RADx Clinical Studies Core team and Test Us At Home Investigators

## Abstract

**Background:** There is a need to understand the performance of rapid antigen tests (Ag-RDT) for detection of the Delta (B.1.61.7; AY.X) and Omicron (B.1.1.529; BA1) SARS-CoV-2 variants.

**Methods:** Participants without any symptoms were enrolled from October 18, 2021 to January 24, 2022 and performed Ag-RDT and RT-PCR tests every 48 hours for 15 days. This study represents a non-pre-specified analysis in which we sought to determine if sensitivity of Ag-RDT differed in participants with Delta compared to Omicron variant. Participants who were positive on RT-PCR on the first day of the testing period were excluded. Delta and Omicron variants were defined based on sequencing and date of first RT-PCR positive result (RT-PCR+). Comparison of Ag-RDT performance between the variants was based on sensitivity, defined as proportion of participants with Ag-RDT+ results in relation to their first RT-PCR+ result, for different duration of testing with rapid Ag-RDT. Subsample analysis was performed based on the result of participants’ second RT-PCR test within 48 hours of the first RT-PCR+ test.

**Results:** From the 7,349 participants enrolled in the parent study, 5,506 met the eligibility criteria for this analysis. A total of 153 participants were RT-PCR+ (61 Delta, 92 Omicron); among this group, 36 (23.5%) tested Ag-RDT+ on the same day, and 84 (54.9%) tested Ag-RDT+ within 48 hours as first RT-PCR+. The differences in sensitivity between variants were not statistically significant (same-day: Delta 16.4% [95% CI: 8.2-28.1] vs Omicron 28.2% [95% CI: 19.4-38.6]; and 48-hours: Delta 45.9% [33.1-59.2] vs. Omicron 60.9% [50.1-70.9]). This trend continued among the 86 participants who had consecutive RT-PCR+ result (48-hour sensitivity: Delta 79.3% [60.3-92.1] vs. Omicron: 89.5% [78.5-96.0]). Conversely, the 38 participants who had an isolated RT-PCR+ remained consistently negative on Ag-RDT, regardless of the variant.

**Conclusions:** The performance of Ag-RDT is not inferior among individuals infected with the SARS-CoV-2 Omicron variant as compared to the Delta variant. The improvement in sensitivity of Ag-RDT noted with serial testing is consistent between Delta and Omicron variant. Performance of Ag-RDT varies based on duration of RT-PCR+ results and more studies are needed to understand the clinical and public health significance of individuals who are RT-PCR+ for less than 48 hours.

## Introduction

Accurate and accessible testing for the SARS-CoV-2 virus is a critical tool for the timely identification of infection to inform isolation recommendations, prevent transmission, and facilitate early initiation of therapy to reduce disease progression.^1^ Rapid antigen tests (Ag-RDT) for COVID-19 show great promise as an easy-to-use, accessible, cost-effective testing modality.^2^ Results from Ag-RDT are available within minutes of sample collection, compared to hours to days for results from reverse-transcription polymerase chain reaction (RT-PCR) tests. The U.S. federal government launched a program in January 2022 to distribute a half billion rapid antigen tests at no cost to U.S. residents in an effort to improve the country’s ability to respond to a surge in the number of COVID-19 cases.^3^

Rapid antigen tests have a lower sensitivity than RT-PCR tests for detecting the SARS-CoV-2 virus;^4^ however, sensitivity can be improved through serial testing.^5^ Existing data on the performance of Ag-RDT predates the emergence of the Omicron (B.1.1.529) variant. The Omicron variant has mutations throughout the SARS-CoV-2 genome; in particular, mutations in the nucleocapsid gene may lead protein conformational changes that affect the target binding site of lateral flow antigen tests. This could theoretically alter performance of Ag-RDT in detecting this variant.^6-9^ The rapid global emergence and dominance of the Omicron variant highlights the importance of understanding the performance of Ag-RDT in the real-world settings.

The urgent need to reassess the performance of Ag-RDT in detecting the Omicron variant is further compounded by early reports that detection of the SARS-CoV-2 Omicron variant by Ag-RDT may be less sensitive than detection of other variants.^10,11^ Recent reports from analytic studies suggest that the Ag-RDT performance does not vary across Delta and Omicron variants.^12-14^ This manuscript presents a comparison of Ag-RDT performance for detection of Delta and Omicron variants of SARS-CoV-2 virus by comparing Ag-RDT to nasal RT-PCR test results when testing participants serially every 48 hours.

## Methods

### Study Population

This analysis used data collected in the Test Us At Home (TUAH) study. TUAH is a prospective cohort study conducted by the NIH Rapid Acceleration of Diagnostics (RADx) program’s Clinical Studies Core and featured a collaboration between the National Institutes of Health, The Food and Drug Administration, and UMass Chan Medical School. Enrollment for the study started on October 18, 2021 and is ongoing. Individuals over the age of 2 years residing in any US state except Hawaii, Alaska, and Arizona were eligible for TUAH, provided they or their parent/guardian had access to a smartphone and were able to receive mail at home. Individuals with COVID-19 symptoms in the 14 days prior to enrollment, a self-reported positive test for COVID-19 in the past three months, no internet access on their smartphone, or currently residing in the correctional justice system were excluded from the study. Study enrollment was self-directed through the study-specific project under the MyDataHelps app (CareEvolution, LLC). Participants provided electronic consent to participate in the study through the app, and individuals under 18 years were required to assent and provide written parental consent. Participants whose first RT-PCR test in the study resulted as ‘Positive’ or ‘Inconclusive’ (*suggesting that one of two targets were detected)* were excluded from this study analysis to allow us to analyze testing performance in context of RT-PCR+ onset (Supp Fig 1). Four populations were defined in this study. Population A included all participants who had a negative RT-PCR during the study prior to a RT-PCR+ and thus allowed us to analyze testing performance in context of RT-PCR+ onset. Populations B, C, and D, were subsets of Population A, defined by the result of the RT-PCR test taken within 48 hours of first RT-PCR+ test: Population B included participants who had a second RT-PCR+ within 48 hours of their first RT-PCR+, Population C included participants who tested RT-PCR-within 48 hours of their first RT-PCR+, and Population D included participants who did not perform another RT-PCR test within 48 hours of the first RT-PCR+ either due to non-adherence or end of the study period (Supp Fig 1). The study protocol for the main study was approved by the Institutional Review Board at UMass Chan Medical School and externally by Western Institutional Review Board.

**Figure 1:**
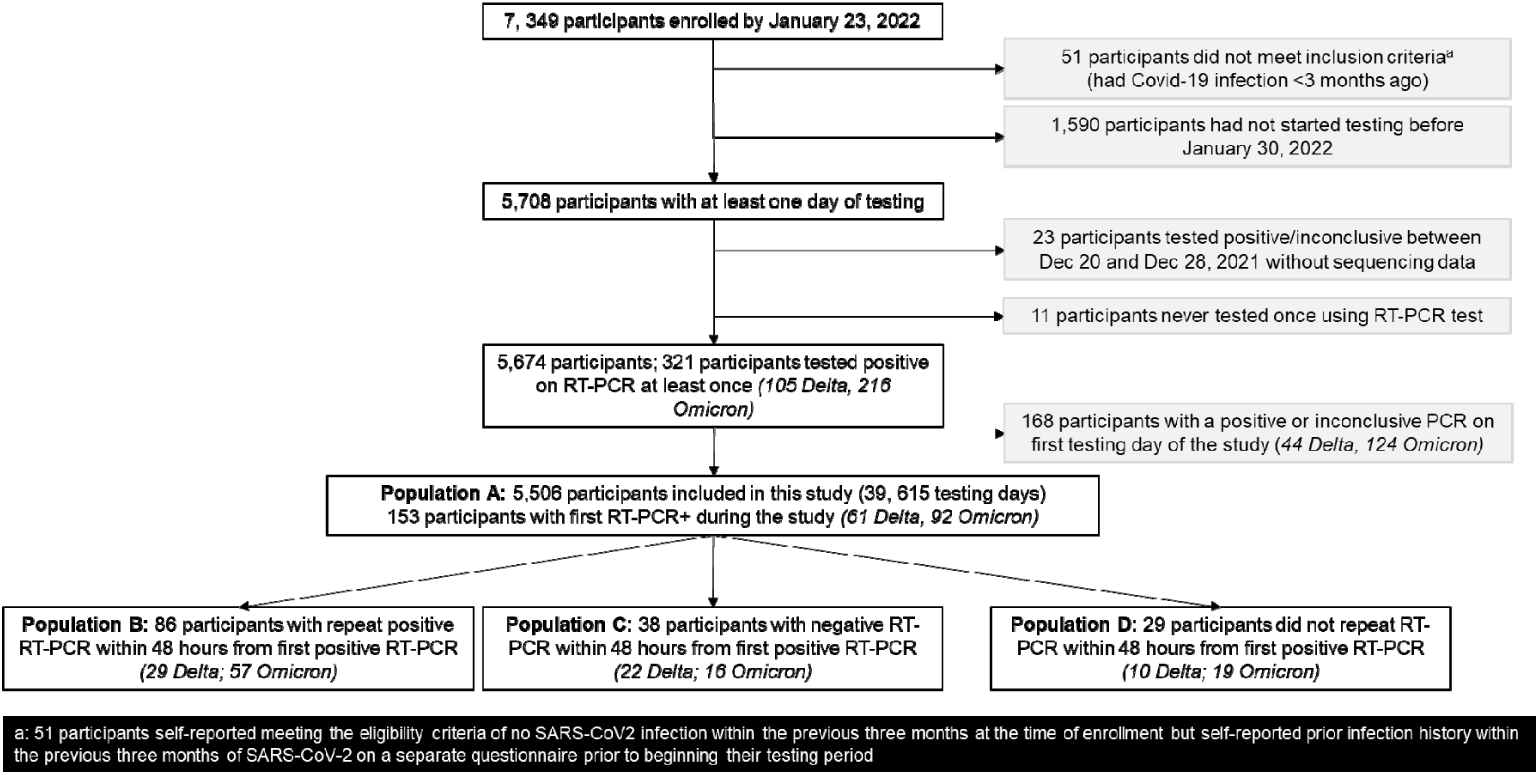
Test Us At Home Participant CONSORT Diagram.

### Study Procedures

Participants were assigned to one of three Emergency Use Authorized (EUA-authorized) Ag-RDT tests on enrollment (BD Veritor™ At-Home COVID-19 Test, Quidel QuickVue At-Home OTC COVID-19 Test, and Abbott BinaxNOW COVID-19 Antigen Self Test). Participants received the Ag-RDT and the EUA-authorized Quest Diagnostics Collection Kit for COVID-19 kits (RT-PCR home-collection kits) by mail to the shipping address provided on enrollment, and a total of 10 Ag-RDT and 7 RT-PCR home-collection kits were provided to participants. Participants were asked to perform the Ag-RDT and self-collect specimen for comparator RT-PCR testing on the same day roughly every 48 hours for 15-days. Participants were instructed to have at least a 15-minute break between the Ag-RDT and the sample collection for RT-PCR tests. On day 15, participants were asked to test only with the Ag-RDT (no PCR testing, Supplemental Table 1). Instructions for performing the tests and for self-collecting and shipping the comparator specimens were provided. All RT-PCR tests were performed at a single CLIA-certified laboratory (Quest Labs, Marlborough, MA) using the Quest Diagnostics RC SARS-CoV-2 assay. This assay is a real-time RT-PCR assay based on the Roche cobas SARS-CoV-2 assay and EUA-authorized for use with specimens collected with the Quest Diagnostics Collection Kit for COVID-19. The Roche cobas was found to have a limit of detection of 1800 NDU/ml when tested using the FDA SARS-CoV-2 reference panel. For participants who tested positive in December and January and had adequate remnant sample, we performed whole genome sequencing of SARS-CoV-2 by amplicon based next generation sequencing on extracted RNA. Viral RNA was extracted from remnant clinical specimens of SARS-CoV-2 positives submitted for real time RT-PCR testing. The viral RNA genome was reverse transcribed to cDNA and RT-PCR amplified (Gene Specific RT-PCR, GSP) in 4 pools using a total of 98 overlapping primer sets. The GSP products were pooled for each sample, diluted, and subjected to barcoding RT-PCR (BCP). During BCP, barcode sequence was added to each specimen. The barcoded samples were purified with Ampure XP beads, Qubit quantified, and normalized. The normalized pool was size selected and sequenced on an Illumina Novaseq sequencer. Consensus viral genome sequence was generated on primer trimmed sequences. Viral specific primer sequences and consensus viral genome sequence generation method were adapted from the ARTIC network.

#### Variables

The result for an Ag-RDT was based on self-report by the participant with the options of ‘Positive’, ‘Negative’, ‘Invalid’, or ‘Do not Know’. The Ag-RDT result was considered as positive only if participant self-reported the result as ‘Positive’. The result for RT-PCR was based on laboratory output as ‘Positive’, ‘Negative’, ‘Inconclusive’, or ‘Test Not Performed’. An ‘Inconclusive’ RT-PCR result was also considered positive for this analysis because it meant that the RT-PCR test detected one of the two targets of the assay. Vaccination history and prior SARS-CoV-2 infection history were based on self-report using the MyDataHelps app. Positive cases were assigned to the Omicron group based on a positive RT-PCR test from a sample collected on January 01, 2022 or later; positive cases were assigned to the Delta group based on a positive RT-PCR test from a sample collected before December 20, 2021. Sequencing results of participants who tested positive in December and the first two weeks of January revealed that the last non-Omicron sample was collected on December 28 and the first participant infected with the Omicron variant was collected on December 20. Participants who tested positive between December 19 and December 31, 2021 were assigned to their respective group based on the sequencing results; those without sequencing results in this period were excluded (Supplemental Table 2 and Figure 1).

#### Analysis

This is not the pre-specified study analysis but was subsequently developed to address an ancillary research question from this unique and comprehensive longitudinal dataset. Specific analysis related to symptomatic status was not pursued due to overlap with the primary objectives of the parent study. Descriptive statistics were performed at the participant level using tabulation of frequencies for categorical data and differences were compared using chi-square or Fisher’s exact test, depending on the cell sample size. We calculated proportions of participants who tested positive on Ag-RDT after their first RT-PCR+ for the different populations described in Figure 1. Numerator was based on participants who had at-least one Ag-RDT+ in the corresponding timeframe since the first RT-PCR+ (same-day, within 48 hours, within 96 hours, within one week). Denominator was based on total number of eligible participants with RT-PCR+ in each population. Corresponding 95% confidence intervals for each proportion were calculated using the exact Clopper-Pearson confidence interval. Additionally, we used a multilevel logistic and logistic regression models to estimate predicted probabilities of Ag-RDT for different cycle threshold (CT) values at the unit of tests and participants, respectively. Specifically, a multilevel generalized linear model with logit link was fitted with Ag-RDT as the dependent variable, and independent variable was an interaction term between variant type (Delta vs Omicron) and RT-PCR CT values (continuous). Predicted probability at 1-unit increments of CT value was estimated using marginal means, and error estimates were calculated using the Delta method that employs Taylor linearization. Similarly, a generalized linear model with logit link was fitted with Ag-RDT positive test within 48 hours of the first RT-PCR+ as the dependent variable, and an interaction term between variant type (Delta vs Omicron) and RT-PCR CT values at the first test (continuous) were modeled. All statistical analyses were performed using Stata.

## Results

### Cohort Characteristics and RT-PCR Test Results

A total of 5,674 participants enrolled in the TUAH study and performed home-based testing between October 21, 2022 and January 29, 2022. This analysis includes data of 5,506 participants after excluding 1) 23 participants because their initial RT-PCR+ was between December 20-28, 2021 and we were unable to obtain sequencing data, 2) 11 participants who never collected a sample for RT-PCR test, and 3) 168 participants who started the study with a RT-PCR+ result on first day of testing (Figure 1). Data from 39,615 days of testing was available from this analytic sample of 5,506 participants. During the study period, 153 participants (61 Delta, 92 Omicron) had an initial RT-PCR+ test and were classified as Population A (Table 1, Figure 1). Of these participants, 86 (56.2%) had a subsequent RT-PCR+ within 48 hours of the first RT-PCR+ test (Population B), 38 (24.8%) had a subsequent RT-PCR− within 48 hours (Population C), and 29 (19.0%) did not have a RT-PCR test 48 hours after their first RT-PCR+ result (Population D). The proportion of individuals with singleton RT-PCR+ results (Population C) was twice as high in participants infected with the Delta variant (36.1%) in comparison to Omicron (17.4%) (p = 0.04). Similar proportions of participants who tested positive on RT-PCR (Population A) were not vaccinated: Delta (21.3%) and Omicron (29.4%) (p = 0.35).

**Table 1:**
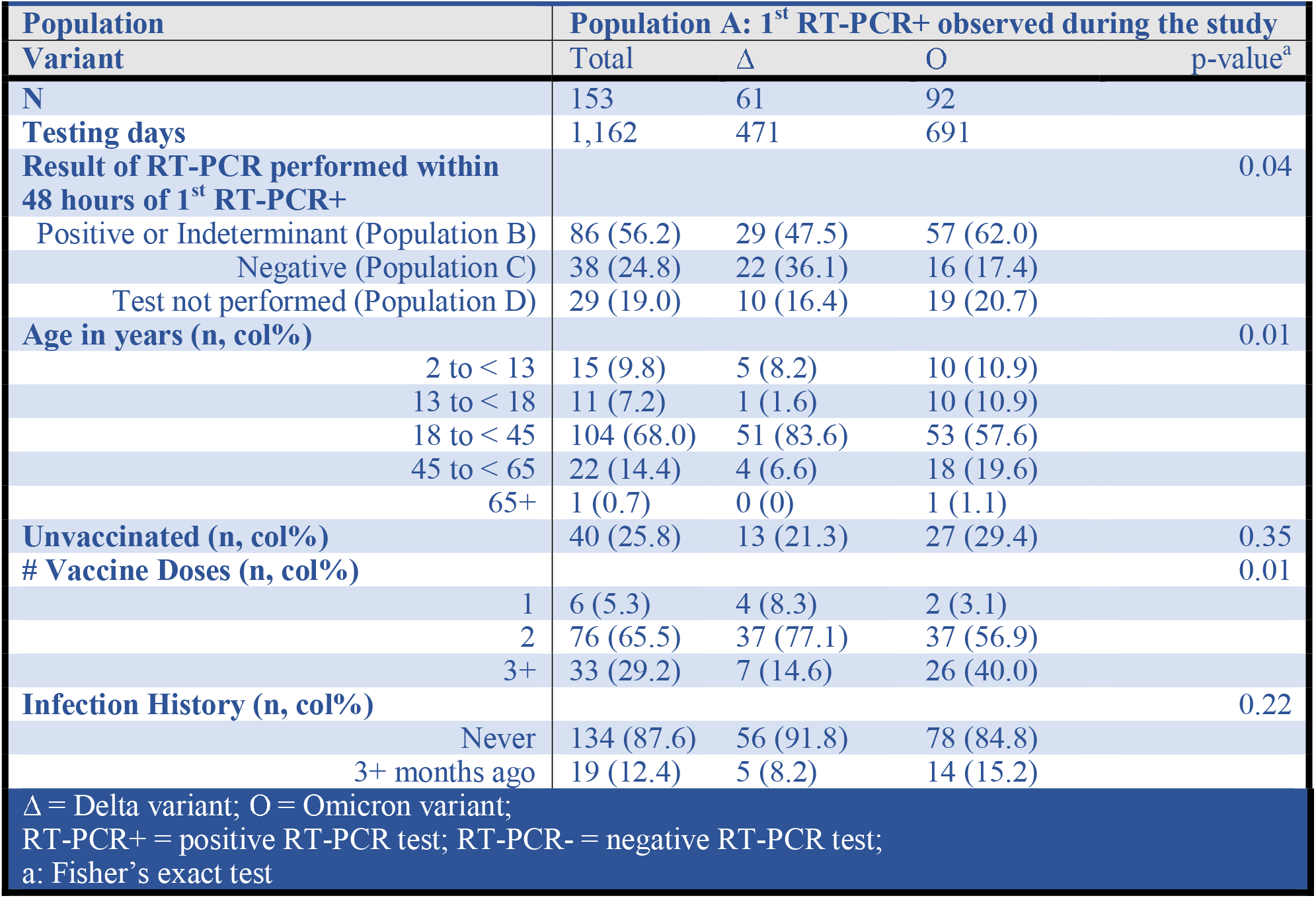
Distribution of participant characteristics based on the variant type.

|  |  |  |  |  |
| --- | --- | --- | --- | --- |
| Population | Population A: 1<sup>st</sup> RT-PCR+ observed during the study |  |  |  |
| Variant | Total | Δ | O | p-value<sup>a</sup> |
| **N** | 153 | 61 | 92 |  |
| **Testing days** | 1,162 | 471 | 691 |  |
| **Result of RT-PCR performed within 48 hours of 1<sup>st</sup> RT-PCR+** |  |  |  | 0.04 |
| Positive or Indeterminant (Population B) | 86 (56.2) | 29 (47.5) | 57 (62.0) |  |
| Negative (Population C) | 38 (24.8) | 22 (36.1) | 16 (17.4) |  |
| Test not performed (Population D) | 29 (19.0) | 10 (16.4) | 19 (20.7) |  |
| **Age in years (n, col%)** |  |  |  | 0.01 |
| 2 to < 13 | 15 (9.8) | 5 (8.2) | 10 (10.9) |  |
| 13 to < 18 | 11 (7.2) | 1 (1.6) | 10 (10.9) |  |
| 18 to < 45 | 104 (68.0) | 51 (83.6) | 53 (57.6) |  |
| 45 to < 65 | 22 (14.4) | 4 (6.6) | 18 (19.6) |  |
| 65+ | 1 (0.7) | 0 (0) | 1 (1.1) |  |
| **Unvaccinated (n, col%)** | 40 (25.8) | 13 (21.3) | 27 (29.4) | 0.35 |
| **# Vaccine Doses (n, col%)** |  |  |  | 0.01 |
| 1 | 6 (5.3) | 4 (8.3) | 2 (3.1) |  |
| 2 | 76 (65.5) | 37 (77.1) | 37 (56.9) |  |
| 3+ | 33 (29.2) | 7 (14.6) | 26 (40.0) |  |
| **Infection History (n, col%)** |  |  |  | 0.22 |
| Never | 134 (87.6) | 56 (91.8) | 78 (84.8) |  |
| 3+ months ago | 19 (12.4) | 5 (8.2) | 14 (15.2) |  |
| Δ = Delta variant; O = Omicron variant;<br>RT-PCR+ = positive RT-PCR test; RT-PCR- = negative RT-PCR test;<br>a: Fisher's exact test |  |  |  |  |

### Time from RT-PCR positivity to Antigen Positivity among Delta and Omicron Variants

Among the 153 participants in Population A whose first RT-PCR+ was observed during the study period, 36 participants tested positive on rapid antigen test (Ag-RDT+) on the same day (23.5%, 95% CI:16.8-30.7) and 84 (54.9, 46.0-62.2%) tested positive within 48 hours from the first RT-PCR+ (Table 2 and Figure 2). The proportions of Omicron-infected participants who were Ag-RDT+ on the same-day, within 48 hours, within 96 hours, and within a week of the first RT-PCR+ result was slightly higher in comparison to Delta infected participants who were Ag-RDT+, but these differences were not statistically significant.

**Table 2:** Distribution of RT-PCR and Ag-RDT test results for different populations used in this analysis.

| Population | A: First RT-PCR+ observed during the study |  |  | B: First RT-PCR+ followed by a 2nd RT-PCR+ in 48 hours |  |  | C: First RT-PCR+ followed by RT-PCR- in 48 hours |  |  | D: Missing 2nd RT-PCR+ in 48 hours from the first RT-PCR+ |  |  |
| --- | --- | --- | --- | --- | --- | --- | --- | --- | --- | --- | --- | --- |
| Variant | Total | Δ | O | Total | Δ | O | Total | Δ | O | Total | Δ | O |
| N | 153 | 61 | 92 | 86 | 29 | 57 | 38 | 22 | 16 | 29 | 10 | 19 |
| Testing days | 1,162 | 471 | 691 | 670 | 231 | 439 | 288 | 170 | 118 | 204 | 70 | 134 |
| Ag-RDT result in comparison to first RT-PCR+ (n, col%) |  |  |  |  |  |  |  |  |  |  |  |  |
| Positive same-day | 36 (23.5) | 10 (16.4) | 26 (28.3) | 31 (36.1) | 8 (27.6) | 23 (40.4) | 0 (0) | 0 (0) | 0 (0) | 5 (17.2) | 2 (20.0) | 3 (15.8) |
| Positive w/in 48hrs | 84 (54.9) | 28 (45.9) | 56 (60.9) | 74 (86.0) | 23 (79.3) | 51 (89.5) | 0 (0) | 0 (0) | 0 (0) | 10 (34.5) | 5 (20.0) | 5 (26.3) |
| Positive w/in 96hrs | 92 (60.1) | 31 (50.8) | 61 (66.3) | 82 (93.5) | 26 (89.7) | 56 (98.2) | 0 (0) | 0 (0) | 0 (0) | 10 (34.5) | 5 (50.0) | 5 (26.3) |
| Positive w/in a week | 93 (60.8) | 31 (50.8) | 62 (67.4) | 83 (96.5) | 26 (89.7) | 57 (100) | 0 (0) | 0 (0) | 0 (0) | 10 (34.5) | 5 (50.0) | 5 (26.3) |
| Negative | 60 (39.2) | 30 (49.2) | 30 (32.6) | 3 (3.5) | 3 (10.3) | 0 (0) | 38 (100) | 22 (100) | 16 (100) | 19 (65.5) | 5 (50.0) | 14 (73.7) |
| Lowest RT-PCR+ CT count (n, col%) |  |  |  |  |  |  |  |  |  |  |  |  |
| 10 to <15 | 4 (2.6) | 3 (4.9) | 1 (1.1) | 3 (3.5) | 2 (6.9) | 1 (1.8) | 0 (0) | 0 (0) | 0 (0) | 0 (0) | 0 (0) | 0 (0) |
| 15 to <19 | 32 (20.9) | 15 (24.6) | 17 (18.5) | 30 (34.9) | 14 (48.3) | 16 (28.1) | 0 (0) | 0 (0) | 0 (0) | 2 (6.9) | 1 (10.0) | 1 (5.3) |
| 20 to <25 | 40 (26.1) | 7 (11.5) | 33 (35.9) | 39 (45.4) | 6 (20.7) | 33 (57.9) | 0 (0) | 0 (0) | 0 (0) | 1 (3.5) | 1 (10.0) | 0 (0) |
| 25 to <30 | 9 (5.9) | 2 (3.3) | 7 (7.6) | 6 (7.0) | 1 (3.5) | 5 (8.8) | 0 (0) | 0 (0) | 0 (0) | 3 (10.3) | 1 (10.0) | 2 (10.5) |
| 30 to <35 | 23 (15.0) | 6 (9.8) | 17 (18.5) | 3 (3.5) | 1 (3.5) | 2 (3.5) | 10 (26.3) | 3 (13.6) | 7 (43.8) | 10 (34.5) | 2 (20.0) | 8 (42.1) |
| 35+ | 8 (5.2) | 4 (6.6) | 4 (4.3) | 0 (0) | 0 (0) | 0 (0) | 6 (15.8) | 4 (18.2) | 2 (12.5) | 2 (6.9) | 0 (0) | 2 (10.5) |
| Missing | 37 (24.2) | 24 (39.3) | 13 (14.1) | 5 (5.8) | 5 (17.2) | 0 (0) | 22 (57.9) | 15 (68.2) | 7 (43.8) | 11 (37.9) | 5 (50.0) | 6 (31.6) |

Δ = Delta variant; O = Omicron variant; RT-PCR+ = positive RT-PCR test; RT-PCR- = negative RT-PCR test; Ag-RDT = Rapid Antigen Test

**Figure 2:**
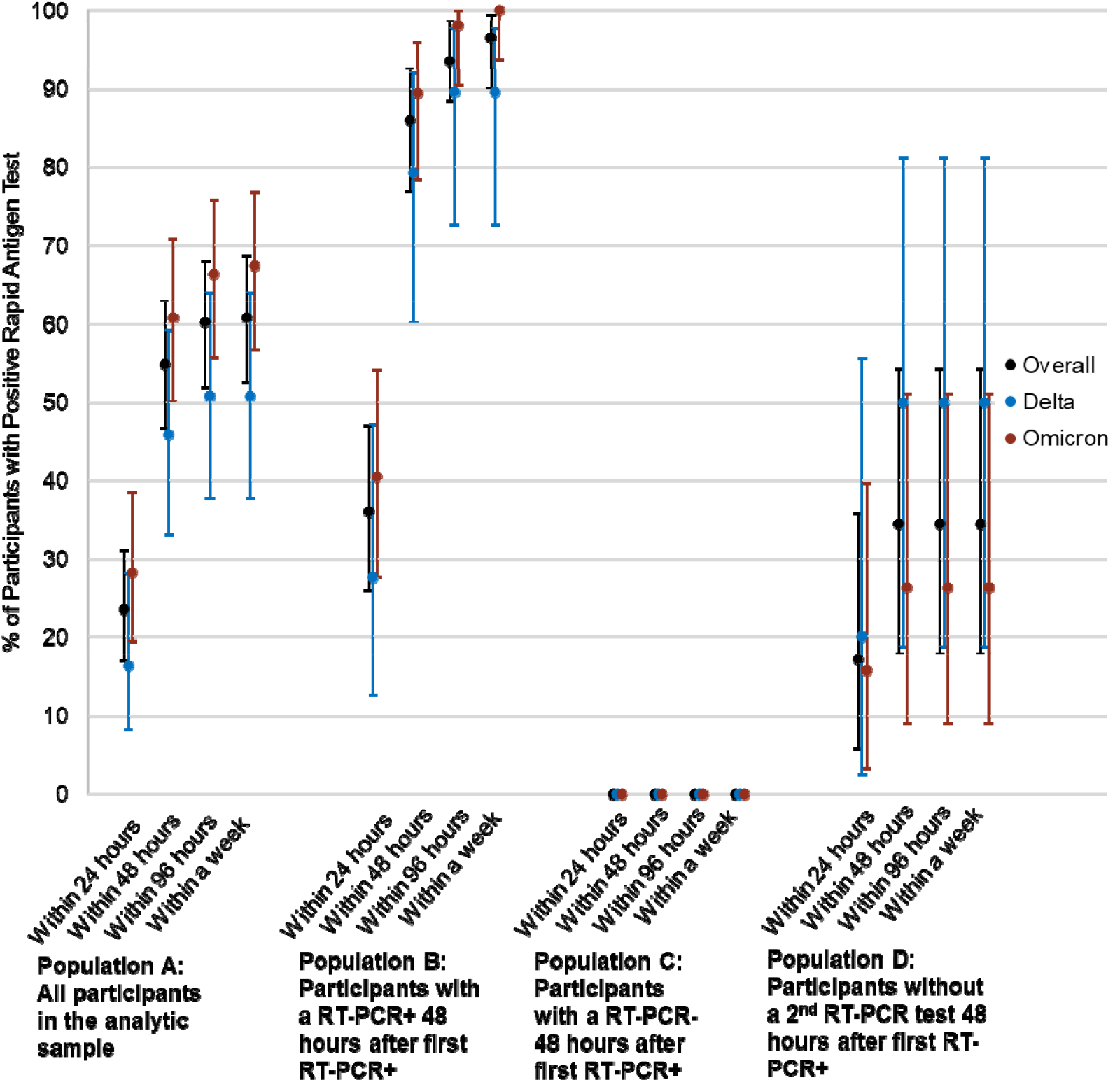
Proportion of participants testing positive by rapid antigen tests by days since initial positive RT-PCR sample collection.

When considering participants with at least 2 sequential RT-PCR+ positive tests (Population B), 86.0% (95% CI: 76.9 – 92.6) of the cases were Ag-RDT+ within 48 hours from first RT-PCR+ result. Of the 86 participants who were serially positive on RT-PCR for at least 48 hours, a higher proportion of Omicron-infected participants tested positive on rapid antigen tests within 48 hours from the first RT-PCR+ compared to Delta-infected participants, but the difference was not statistically significant (Delta 79.3% [95% CI: 60.3-92.1] vs Omicron 89.5% [78.5-96.0]) (Figure 2). The proportion of participants who never tested positive on Ag-RDT+ differed significantly across the different populations (Population A: 39.2% [31.4 – 47.4] vs. Population B: 3.5% [0.7-9.1%]) and ***all*** 38 participants in Population C never had a positive rapid antigen test during the study. Of note, none of the Population C participants had a CT value lower than 30 (Table 2).

### Relationship between probability of Ag-RDT positivity and CT value among Delta and Omicron Variants

We observed similar sensitivity between variants for same-day positivity of Ag-RDT tests in comparison to RT-PCR+ when CT count was less than 30 (Delta: 83.3% vs Omicron: 87.2%) and for 48-hour positivity of Ag-RDT in comparison to RT-PCR+ when CT count was less than 30 (Delta: 90.9% vs Omicron: 95.8%). The corresponding predicted probabilities for these scenarios were calculated based on these data and shown in Figure 3A and 3B, respectively. Compared to participants infected with the Delta variant, we observed a higher predicted probability of Ag-RDT+ among Omicron variant when the CT value was lower than 30; however, this difference was not statistically significant (Figure 3A). Results of predicted probability for Ag-RDT+ within 48 hours of first RT-PCR+ as a function of first positive RT-PCR+ CT values is shown in Figure 3B and is based on data from the 100 participants whose CT value from the first RT-PCR+ was available. We observed a higher predicted probability of Ag-RDT+ within 48 hours of first RT-PCR+ when the first positive RT-PCR had a CT value of 34 or lower for individuals with the Omicron variants in comparison to the Delta variant, though the differences were within the margin of error.

**Figure 3:**
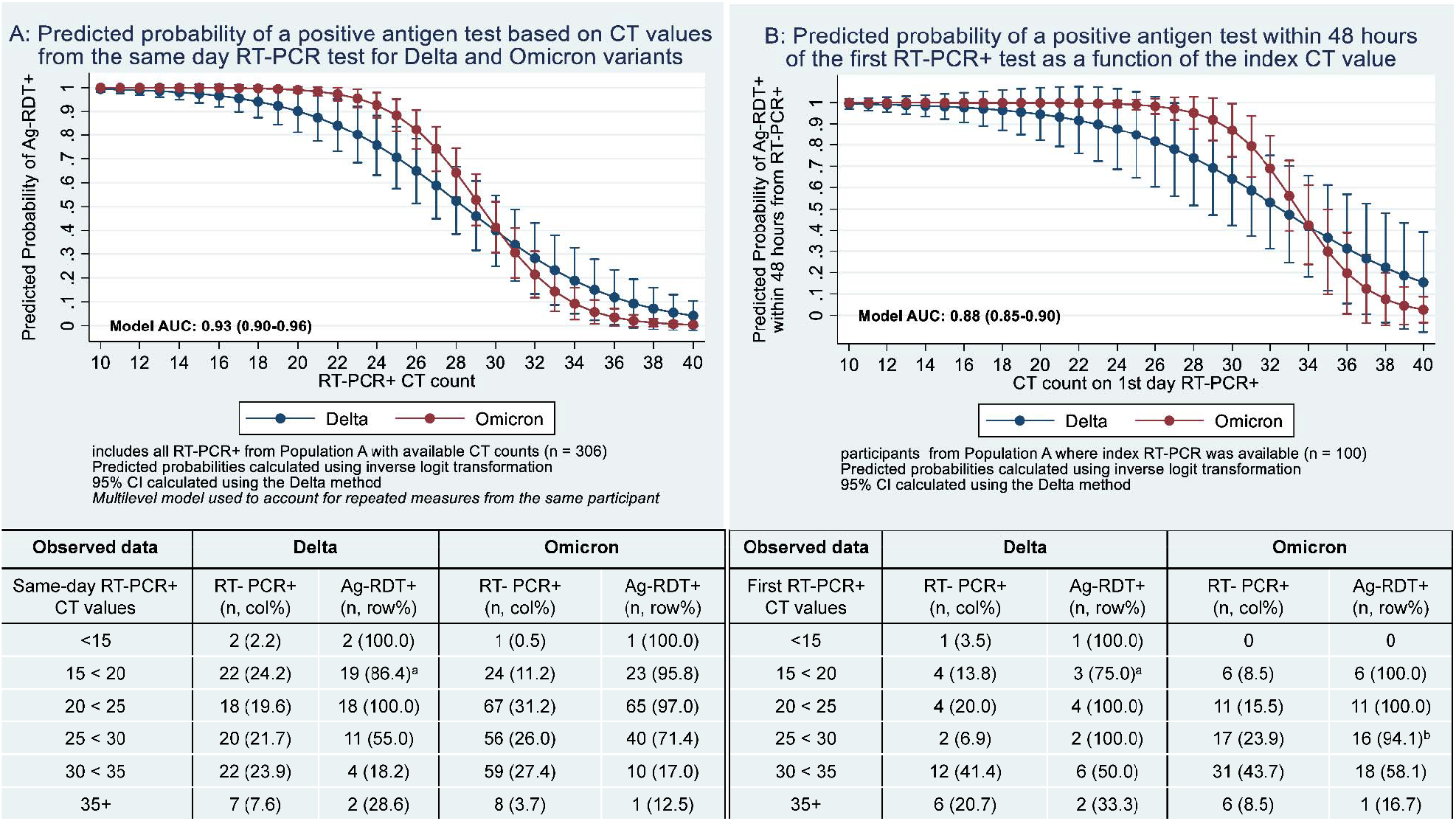
Probability of Antigen Test Positivity of Delta and Omicron Variants as a Function of CT Values Among RT-PCR Positive Participants.

## Discussion

In this analysis of data from 5,506 participants that included 39, 615 days of Ag-RDT and RT-PCR testing spanning October 2021 to January 2022, we found that the performance of Ag-RDT was not inferior for detection of the Omicron variant in comparison to the Delta variant. Although not statistically significant, we found that a higher proportion of participants with Omicron variant had a positive Ag-RDT+ in comparison to participants with Delta variant on the same-day and within 48 hours as RT-PCR+. It is important to note that overall (Population A), the same-day positivity for Ag-RDT on onset of RT-PCR+ was low at 23.5%. Repeated Ag-RDT testing within 48 hours improved this proportion to 54.9% overall. However, among the participants who were positive on RT-PCR test for at least 48 hours, Ag-RDT was positive for 36.1% of the participants on the same day and 86.0% of the participants within 48 hours. The phenomenon of “singleton RT-PCR+” (Population C), where a participant’s first RT-PCR+ result is followed by a negative RT-PCR result within 48 hours, merits further discussion because Ag-RDT tests completely failed to detect the infection, regardless of the variant. Taken together, our findings suggest that Ag-RDT tests detect infections similarly for Delta and Omicron variants, with overall low detection rates on the same day as an initial RT-PCR+ and a higher detection rate when a second test is used 48 hours after the first.

It is important to consider the following factors when interpreting these results: 1) these results present a comparison between Ag-RDT and RT-PCR tests using self-collected nasal swabs; 2) the testing frequency of 48 hours does not allow a finer temporal resolution of the analysis of test performance; and 3) the results of Ag-RDT are based on self-report. However, these limitations are non-differential and unlikely to bias the comparison of Ag-RDT performance between Delta and Omicron variants. Furthermore, the data collected from this study illustrates performance of Ag-RDT self-collected at home, which is a different setting than proctored or on-site administration of Ag-RDT with guidance from trained individuals or when tests are performed by health care professionals.

Early epidemiologic studies have shown decreased and delayed sensitivity of Ag-RDT in detecting the Omicron variant in comparison to saliva RT-PCR testing. Notably, in a pre-print, Adamson et al. reported that among 28 people with a positive saliva RT-PCR test with suspected Omicron variant infection and CT value < 29, none tested positive on nasal Ag-RDT within 24 hours.^10^ Additionally, they reported a median lag of 3 days from saliva RT-PCR positivity to a positive Ag-RDT from nasal swab. By contrast, among the infections where initial nasal RT-PCR+ CT value was lower than 30, our study found that Ag-RDT was positive within 48 hours in 33 out of the 34 instances. In the sole case where Ag-RDT+ was not recorded within 24 hours of the initial RT-PCR+ with a CT value < 30, a subsequent Ag-RDT was not performed. We also found a sensitivity of 87.2% with Ag-RDT performed on same-day as RT-PCR+ with CT count < 30 among individuals infected with the Omicron variant, which is similar to a separate report that evaluated similar performance among predominantly symptomatic participants.^15^ The discrepancy of findings from our study in comparison the findings by Adamson et al. may be explained by the use of saliva RT-PCR instead of nasal RT-PCR as the primary comparator. Marais et al. reported that the positive percent agreement of RT-PCR tests from saliva and mid-turbinate swab compared to a composite standard for the Delta variant was 71% (95% CI: 53-84%) and 100% (95% CI: 89-100%), respectively, but 100% (95% CI: 90-100%) and 86% (95% CI: 71-94%) for the Omicron variant.^16^ However, considering that saliva PCR tests are not widely available and the typical turn-around-time for commercial nasal RT-PCR tests ranges from 36-48 hours, our finding suggests that serial use of Ag-RDT may be a viable option for ascertaining infection status of SARS-CoV-2 infection, regardless of Delta or Omicron variant.

The findings from our study reinforce the importance of serial use of Ag-RDTs to overcome the relatively low sensitivity of Ag-RDT on the first-day of RT-PCR positivity, which is consistent with previous findings by Smith et al.^5^ In that study of known positives and close-contacts, limited sensitivity was observed for a single time-point Ag-RDT in the early course of infection, but repeated testing every 48 hours or every 72 hours improved sensitivity from lower than 40% to higher than 80%. There is a suggestion that viral dynamics with Omicron infection may be different, such that there is a more rapid rise in the RNA viral load but lower peak and a shorter clearance phase in comparison to the Delta variant.^17^ Indeed, we observed a slightly higher proportion of first CT values less than 30 for Omicron infections (34/71, 47.8%) than for Delta infections (11/29, 37.9%). Our findings of higher first-day sensitivity with Ag-RDT among the participants infected with Omicron variant may be attributable to these differences, which were not statistically significant.

In this study, we also observed that nearly half (45.1%) of the participants with a positive RT-PCR test had a false negative result on Ag-RDT even when two antigen tests were performed within 48 hours of the first RT-PCR positivity. However, when restricting the analysis to participants who tested positive on RT-PCR for at least 48 hours (Population B), the false negative rate for Ag-RDT was 14.0% within 48 hours with no significant differences between the variant types. For the population of participants with singleton RT-PCR+, additional studies are needed to understand this phenomenon further in the context of SARS-CoV-2 infection compared to other viral infections where “blips” are commonly described.^18-20^ Factors such as SARS-CoV-2 immune status, local or systemic viral load, or assay limit of detection may play a role. The public health implications of false negative Ag-RDT associated with the singleton RT-PCR+ remains unclear.^21^ Because there is no way to prospectively determine who will remain positive on RT-PCR and who will have a singleton RT-PCR+, it is important to elucidate the significance of our finding that Ag-RDT fail to detect singleton RT-PCR+.

This analysis offers a unique look at longitudinal RT-PCR and Ag-RDT in a large prospective cohort, allowing us to capture data at the onset of infection and during the infection course throughout the emergence of the Omicron variant. This study uses three different Ag-RDTs, which increases generalizability but does not guarantee it, and further evaluation of other Ag-RDTs may be needed as a clinical study. Current identification of variants as Omicron or Delta in this study is based on sequencing of a subset of samples during the month of December and first week of January, instead of all participants who tested positive. However, our observed sequencing results during December and January closely resembles that of CDC’s variant surveillance. To decrease possible misclassification of Delta and Omicron samples, positive participants without sequencing results in the time when both Delta and Omicron variants were circulating were excluded. Furthermore, correction of possible misclassification error is unlikely to reverse the findings that Ag-RDT have equivalent performance for Delta and Omicron variants.

## Conclusion

Self-collected nasal swab Ag-RDT performance was similar between Omicron and Delta variants. In both cases, detection of virus with Ag-RDT was associated with relative viral load as measured by CT value. Our data suggests that the performance of Ag-RDTs remains stable during the Omicron period as compared to the Delta period, and that serial testing continues to be important to raise the performance of Ag-RDTs. Future work to increase our understanding of individuals with singleton RT-PCR positive is needed to determine the public health significance of a false negative Ag-RDT in this subpopulation.

## Data Availability

All data produced in the present study are available upon reasonable request to the authors.

## Acknowledgment

We are deeply grateful first to our many study participants and second to our collaborators from the National Institute of Health (NIBIB and NHLBI) who provided scientific input into the design of this study and interpretation of our results, but could not formally join as co-authors due to institutional policies and to the Food and Drug Administration (Office of In Vitro Diagnostics and Radiological Health) for their involvement in the primary TUAH study. We received meaningful contributions from Drs. Bruce Tromberg, Jill Heemskerk, Felicia Qashu, Dennis Buxton, Erin Iturriaga, Jue Chen, Rachael Fleurence, Andrew Weitz, and Krishna Juluru. Additionally, Quest Diagnostics, LLC provided invaluable support to facilitate the conduct of this study. In particular, we are grateful to Ms. Lisa Cashman, Mr. Scott Burlingame, and Drs. Lokinendi Rao and Karthik Kuppuswamy for their collaboration. Finally, we are thankful to county health departments across the country who helped spread awareness of this study to their constituents; their outreach made the digital siteless approach of this study, a reality.

## Tables and Figures

**Supplemental Table 1:**
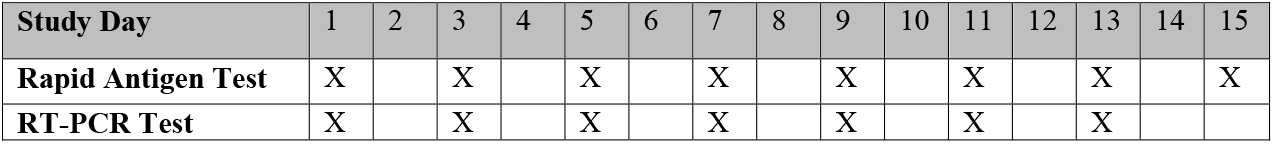
Test Us At Home RT-PCR and Rapid Antigen Testing Schedule.

| Study Day | 1 | 2 | 3 | 4 | 5 | 6 | 7 | 8 | 9 | 10 | 11 | 12 | 13 | 14 | 15 |
| --- | --- | --- | --- | --- | --- | --- | --- | --- | --- | --- | --- | --- | --- | --- | --- |
| **Rapid Antigen Test** | X |  | X |  | X |  | X |  | X |  | X |  | X |  | X |
| **RT-PCR Test** | X |  | X |  | X |  | X |  | X |  | X |  | X |  |  |

**Supplemental Table 2:** Sequencing Results of Positive RT-PCR Samples.

| Sample Collection Date | Number of Delta Sequences Reported* | Number of Omicron Sequences Reported (BA.1) |
| --- | --- | --- |
| 11/22/2021 | 1 | 0 |
| 11/24/2021 | 1 | 0 |
| 11/26/2021 | 2 | 0 |
| 11/29/2021 | 1 | 0 |
| 12/1/2021 | 2 | 0 |
| 12/2/2021 | 1 | 0 |
| 12/3/2021 | 1 | 0 |
| 12/5/2021 | 2 | 0 |
| 12/6/2021 | 4 | 0 |
| 12/7/2021 | 1 | 0 |
| 12/9/2021 | 1 | 0 |
| 12/10/2021 | 2 | 0 |
| 12/11/2021 | 2 | 0 |
| 12/13/2021 | 1 | 0 |
| 12/15/2021 | 1 | 0 |
| 12/16/2021 | 1 | 0 |
| 12/17/2021 | 1 | 0 |
| 12/18/2021 | 2 | 0 |
| 12/19/2021 | 1 | 0 |
| 12/20/2021 | 1 | 1 |
| 12/21/2021 | 2 | 0 |
| 12/22/2021 | 1 | 0 |
| 12/23/2021 | 1 | 0 |
| 12/24/2021 | 1 | 1 |
| 12/27/2021 | 1 | 1 |
| 12/28/2021 | 2 | 1 |
| 1/1/2022 | 0 | 1 |
| 1/4/2022 | 0 | 6 |
| 1/5/2022 | 0 | 1 |
| 1/7/2022 | 0 | 1 |
| 1/8/2022 | 0 | 1 |
| 1/9/2022 | 0 | 1 |
| 1/10/2022 | 0 | 9 |
| 1/11/2022 | 0 | 2 |
| 1/12/2022 | 0 | 1 |
| 1/13/2022 | 0 | 2 |
| 1/14/2022 | 0 | 4 |
| 1/15/2022 | 0 | 3 |
| 1/16/2022 | 0 | 8 |
| 1/17/2022 | 0 | 2 |
| 1/18/2022 | 0 | 1 |
| *Delta sequences were of the lineages AY.100, AY.103, AY.111, AY.113, AY.119, AY.25, AY.25.1, AY.3, AY3.1, AY.33, AY.39, AY.44 |  |  |

